# Estimating the maximum daily number of incident COVID-19 cases manageable by a healthcare system

**DOI:** 10.1101/2020.03.25.20043711

**Authors:** Vasily Giannakeas, Deepit Bhatia, Matthew T. Warkentin, Isaac I. Bogoch, Nathan M. Stall

## Abstract

The COVID-19 Acute and Intense Resource Tool (CAIC-RT) is an interactive online tool capable of estimating the maximum daily number of incident COVID- 19 cases that a healthcare system could manage given age-based case distribution and severity.

## Background

The Coronavirus Disease 2019 (COVID-19) pandemic originated in China in late 2019 and continues to spread globally.^1^ At the time of writing, there were over 260,000 COVID-19 cases causing close to 12,000 deaths across more than 185 affected countries and territories.^2^ As healthcare systems like those in Northern Italy approach collapse, there is a pressing need for tools modeling the capacity of acute and critical care systems during the COVID-19 pandemic.^3^

## Objective

To develop an online tool capable of estimating the maximum daily number of incident COVID- 19 cases that a healthcare system could manage given age-based case distribution and severity.

## Methods

We modeled steady-state patient-flow dynamics for acute care beds, critical care beds, and mechanical ventilators during the COVID-19 pandemic.^4^ Parameters for patient-flow dynamics were extracted from evolving data on COVID-19 and assumptions based on expert guidance, but were left modifiable in the tool for users to adjust based upon local experience. We used the package *shiny* within R (version 3.5.3) to create the interactive tool.

The tool determines the maximum daily number of incident COVID-19 cases (“patients in”) which would equal the maximum daily turnover of acute care beds, critical care beds, and mechanical ventilators available and/or used for COVID-19 patients (“patients out”)—this is the steady state which could be maximally managed by a healthcare system. Resources available for COVID-19 patients include the proportion of existing resources that could be made maximally available to support COVID-19 patients plus any additional surge capacity.

The tool first calculates the daily turnover of acute care beds, critical care beds and mechanical ventilators available for COVID-19 patients by taking the maximally available number of those resources and dividing it by the expected duration of use for COVID-19 patients. Based on published data, the average length of stay in acute care was set at 11 days, the average length of stay in critical care was set at 20 days, and the average length of time for mechanical ventilation was set at 20 days.^1,5^ The tool then calculates the population-weighted expected probabilities of acute care hospitalization and critical care admissions for COVID-19 infections; we assumed that 50% of critical care patients would require mechanical ventilation.^5,6^ Finally, the maximum daily number of incident COVID-19 cases that a healthcare system could manage is calculated by dividing the daily turnover of maximally available acute care beds, critical care beds or mechanical ventilators by the probability of those resources being used among COVID-19 cases.

While the tool can be used in any region, default parameters for age-based case distribution and severity were set using data from the United States, while parameters for acute and critical care resource availability were set for Canada’s most populous province of Ontario (see *Appendix 1*).^6^

## Findings

The COVID-19 Acute and Intensive Care Resource Tool (CAIC-RT) is open-access and published online, available at https://caic-rt.shinyapps.io/CAIC-RT. As a demonstration, the maximum daily number of incident COVID-19 cases which could be managed by the Ontario healthcare system is detailed in *Figure 1*.

**Figure 1:**
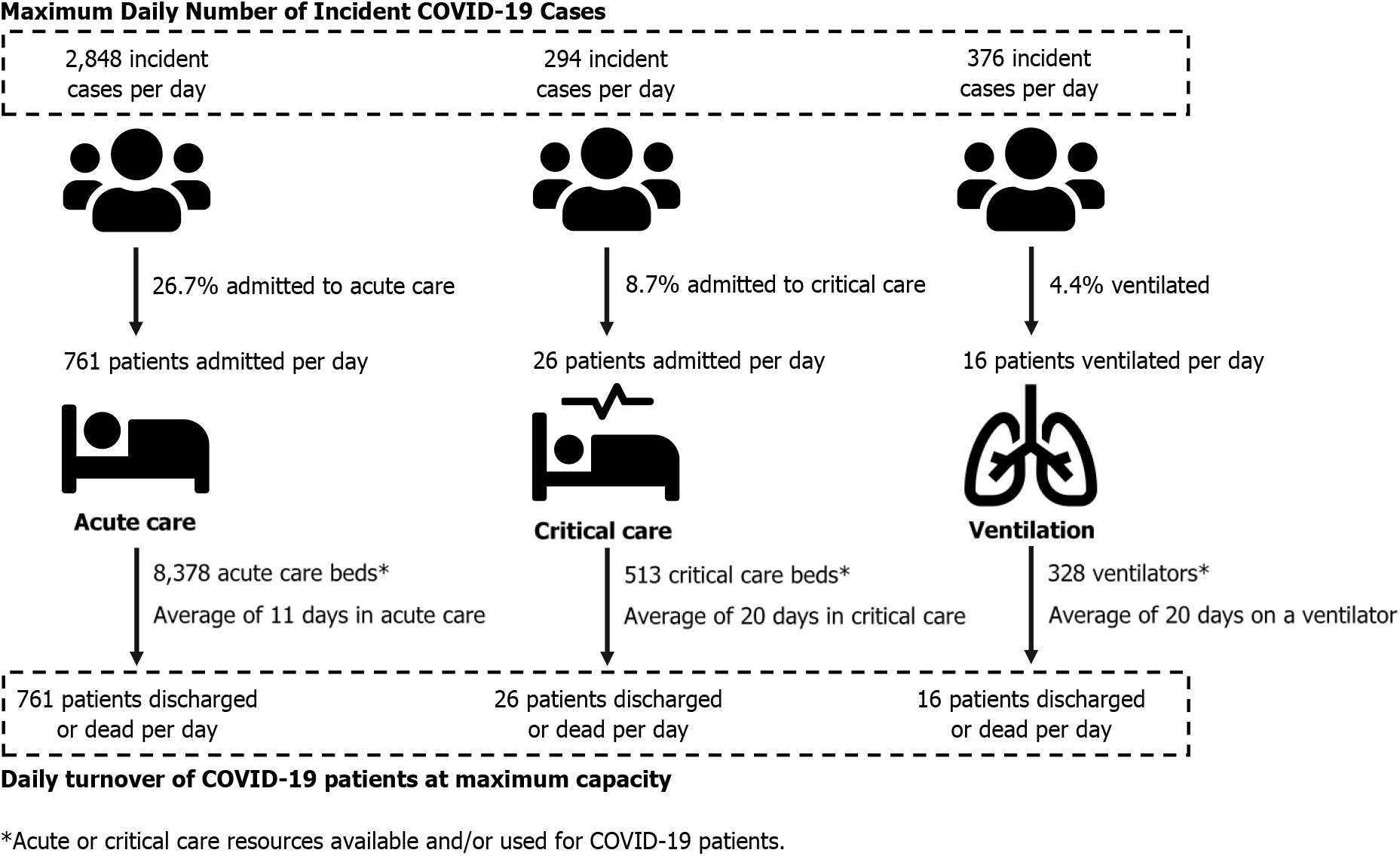
Maximum daily number of incident COVID-19 cases which could be managed by the Ontario healthcare system.

## Discussion

Using an online tool, healthcare systems can estimate the maximum daily number of incident COVID-19 cases which could be managed based on age-based case distribution and severity and the number of maximally available acute and critical care resources. Unlike forecasting tools, our tool has the advantage of determining a sustainable threshold for resource utilization during the COVID-19 pandemic rather than forecasting when resources might become depleted based on assumptions about reporting, epidemic growth and reproduction numbers.

Outputs from the tool will allow planners to examine how increases in acute and critical resources available for COVID-19 patients can impact healthcare system sustainability. Further, the tool can inform the required intensity for non-pharmaceutical societal interventions like physical distancing based on a healthcare system’s proximity to the sustainable threshold. Finally, the tool allows for customization of age-based case distribution and severity which is essential for countries with differing population demographics and healthcare systems.

Limitations of this tool include the assumption that COVID-19 cases become instantaneously hospitalized and the application of Canadian, Chinese and US data for default parameters which may not generalizable to all healthcare systems. Though we intentionally left the tool modifiable, we will update the default values as new data emerges, in order to account for the ramp-up of diagnostic testing in countries like the United States with the understanding that most of those tested will not be hospitalized.

## Data Availability

Study protocol and statistical code are described in the methods and available in Appendix 1 and 2. Data set: N/A.

https://caic-rt.shinyapps.io/CAIC-RT/

## Appendix 1 Parameters for COVID-19 Acute and Intensive Care Resource Tool (CAIC-RT)

### 1. Age-Based Case Distribution and Severity

**TABLE.** Hospitalization, intensive care unit (ICU) admission, and case–fatality percentages for reported COVID–19 cases, by age group — United States, February 12–March 16,2020

| Age group (yrs)<br>(no. of cases) | %* |  |  |
| --- | --- | --- | --- |
|  | Hospitalization | ICU admission | Case-fatality |
| 0–19 (123) | 1.6–2.5 | 0 | 0 |
| 20–44 (705) | 14.3–20.8 | 2.0–4.2 | 0.1–0.2 |
| 45–54 (429) | 21.2–28.3 | 5.4–10.4 | 0.5–0.8 |
| 55–64 (429) | 20.5–30.1 | 4.7–11.2 | 1.4–2.6 |
| 65–74 (409) | 28.6–43.5 | 8.1–18.8 | 2.7–4.9 |
| 75–84 (210) | 30.5–58.7 | 10.5–31.0 | 4.3–10.5 |
| ≥85 (144) | 31.3–70.3 | 6.3–29.0 | 10.4–27.3 |
| **Total (2,449)** | **20.7–31.4** | **4.9–11.5** | **1.8–3.4** |

\* Lower bound of range = number of persons hospitalized, admitted to ICU, or who died among total in age group; upper bound of range = number of persons hospitalized, admitted to ICU, or who died among total in age group with known hospitalization status, ICU admission status, or death.

The age-based case distribution and severity parameters took the midpoint of the lower and upper bound of ranges for hospitalization and ICU admission in the above table.

*Source:* CDC COVID-19 Response Team. Severe Outcomes Among Patients with Coronavirus Disease 2019 (COVID-19) — United States, February 12–March 16, 2020. MMWR Morb Mortal Wkly Rep. 2020.

### 2. Average Acute and Critical Care Resource Utilization by COVID-19 Patients

| Resource | Value | Source(s) |
| --- | --- | --- |
| Length of stay, acute care wards (excluding time spent in critical care) | 11 days | Zhou F, Yu T, Du R, et al. Clinical course and risk factors for mortality of adult inpatients with COVID-19 in Wuhan, China: a retrospective cohort study. *Lancet.* 2020. |
| Length of stay, critical care (excluding time spent on acute care wards) | 8-20 days | Zhou F, Yu T, Du R, et al. Clinical course and risk factors for mortality of adult inpatients with COVID-19 in Wuhan, China: a retrospective cohort study. *Lancet.* 2020.<br>Yang X, Yu Y, Xu J, Shu H, Xia J, Liu H, et al. Clinical course and outcomes of critically ill patients with SARS-CoV-2 pneumonia in Wuhan, China: a single-centered, retrospective, observational study. *Lancet Respir Med.* 2020. |
| Length of time, mechanical ventilation | 7.5-20 days | Wang D, Hu B, Hu C, et al. Clinical Characteristics of 138 Hospitalized Patients With 2019 Novel Coronavirus-Infected Pneumonia in Wuhan, China. *JAMA.* 2020.<br>Yang X, Yu Y, Xu J, Shu H, Xia J, Liu H, et al. Clinical course and outcomes of critically ill patients with SARS-CoV-2 pneumonia in Wuhan, China: a single-centered, retrospective, observational study. *Lancet Respir Med.* 2020. |

### 3. Estimated Acute and Critical Care Resources Maximally Available for COVID-19 Patients Province of Ontario, Canada

In Canada’s most populous province of Ontario with nearly 15-million residents, there are approximately 33,511 acute care beds, 2053 ICU beds, and 1311 mechanical ventilators. Assuming 75% baseline occupancy rates of these resources for non-COVID-19 patients, there are **8,378 available acute care beds, 513 available ICU beds, and 328 available mechanical ventilators**.

*Source:* Barrett K, Khan Y, Mac S, Ximenes R, Naimark D, Tuite A, et al. COVID-19: Predicting Healthcare Resource Needs in Ontario 2020 [updated March 18, 2020. Available from: https://drive.google.com/drive/folders/1T5I2VKuvVFD0FmFGItcH_ZX1XioH4vYU.

#### United States

The *Society of Critical Care Medicine* (SCCM) Ventilator Taskforce has estimated acute and critical care resource availability for the COVID-19 pandemic in the United States. Based on 2018 data from the American Hospital Association, there are approximately 438,368 staffed acute care beds, 96,596 ICU beds and 62,000 full-featured mechanical ventilators. The taskforce assumes 66% baseline occupancy rates of these resources for non-COVID-19 patients, which yields **144,661 available acute care beds, 31,877 available ICU beds, and 20,460 available mechanical ventilators**.

*Source:* Society of Critical Care Medicine Ventilator Taskforce. U.S. ICU Resource Availability for COVID-19. https://sccm.org/getattachment/Blog/March-2020/United-States-Resource-Availability-for-COVID-19/United-States-Resource-Availability-for-COVID-19.pdf?lang=en-US. Published 2020. Updated March 19, 2020. Accessed March 19, 2020.

## Appendix 2 Technical Appendix

We determined the maximum daily number of incident COVID-19 cases which could be managed by a healthcare system. The tool uses steady state analysis and determines the maximum daily number of COVID-19 cases (“patients in”) which would equal the maximum daily turnover of acute care beds, critical care beds, and mechanical ventilators (“patients out”)—this represents the steady state which could be maximally managed by a healthcare system.

**Appendix 2 Figure 1:**
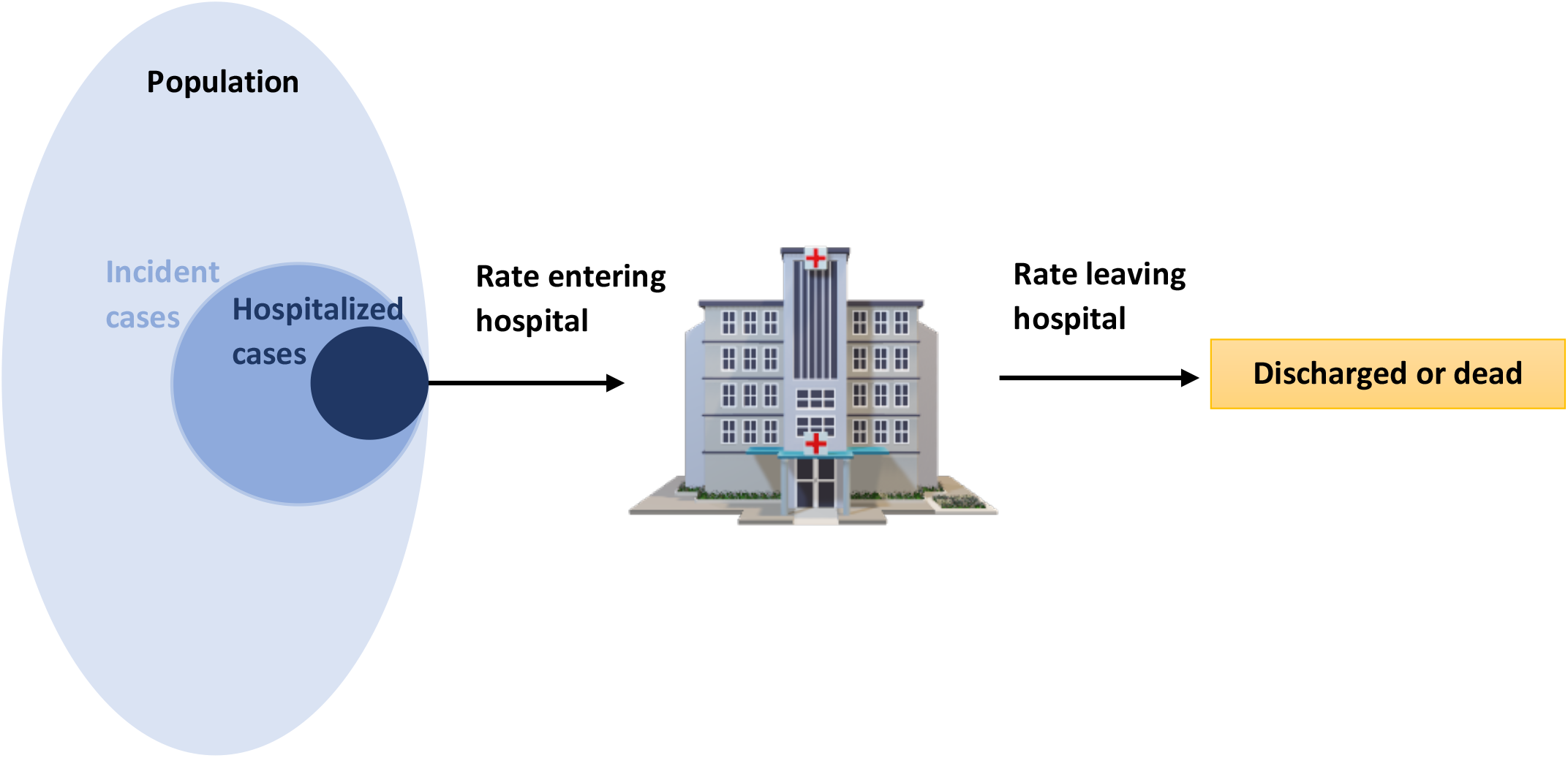
Flowchart of incident COVID-19 cases entering and exiting a hospital.

In the example above we can see that within a given population there are incident cases (new COVID-19 cases). Some of these cases are hospitalized and enter the hospital. COVID-19 cases that are admitted leave the hospital either through discharge or death. The goal is to determine the maximum rate of incident cases that can occur in the population such that, given the hospital’s maximum capacity, the rate of COVID-19 patients requiring hospitalization is equal to the rate of COVID-19 patients leaving the hospital.

### Step 1: Calculate the rate of cases leaving the hospital when at capacity

We calculate the rate of patients leaving the hospital by taking the number of COVID-19 patients at the hospital when at capacity by the ***average*** length-of-stay a patient spends at the hospital. It is important to note that the ***average/mean*** length-of-stay is to be used and not the median length-of-stay. This is referred to as the sojourn time and represents the turnover rate that occurs within hospital.

For example, suppose the hospital can handle 100 patients, 50% of beds are usually occupied by non-COVID-19 patients and 50% are available for or occupied by COVID-19 patients. Further, suppose the average length-of-stay for COVID-19 patients is 10 days. We can calculate the rate of COVID-19 patients leaving the hospital as:

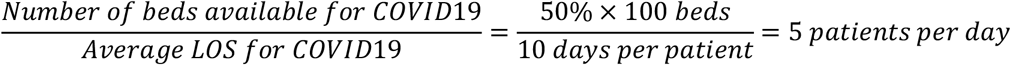

Therefore, given that there are 50 beds available for COVID-19 patients, and each patient stays 10 days on average, the expected rate of COVID-19 patients leaving the hospital is 5 patients per day.

### Step 2: Calculate the proportion of COVID-19 cases that get hospitalized

The next step is to determine the proportion of COVID-19 patients that become hospitalized. Referring to *Appendix 2 Figure 1* we determined what proportion of the incident cases (mid-blue circle) are hospitalized (dark-blue circle). The probability of being hospitalized given someone is a COVID-19 case is highly dependent on the age distribution of the case population. To perform this calculation, among cases we multiply the probability of being within an age-group by the probability of being hospitalized if someone is in that age group. The sum of all age-groups gives the overall probability of being hospitalized per case:

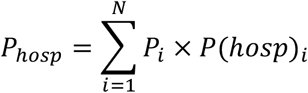

*P*_*hosp*_: Expected probability of being hospitalized given someone is a case

*P*_*i*_: Probability of being within age-group : *i* If someone is a case, where 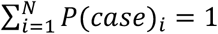

*P(hosp)*_*i*_: Probability of being hospitalized given someone is a case in age-group *i*

*N*: Number of unique age-groups

For example, suppose the case population distribution and probability of hospitalization was as follows:

| Age-group | Proportion of cases [%] | Probability of hospitalization [%] |
| --- | --- | --- |
| <50 | 10% | 5% |
| 50-69 | 30% | 10% |
| 70-79 | 20% | 15% |
| 80+ | 40% | 40% |

The expected probability of hospitalization in this population will be:

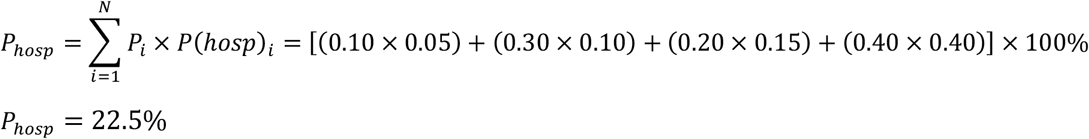

Therefore, we expect 22.5% of the cases in this population to be hospitalized.

### Step 3: Calculate the maximum daily number of COVID-19 cases sustainable by a healthcare system

The final step is to calculate the maximum daily cases of COVID-19 that can occur within the population such that the hospital can run at full-capacity with no patient over-flow (i.e. steady-state at full capacity). Given what we calculated in Step 1 and Step 2, for the system to be steady-state:

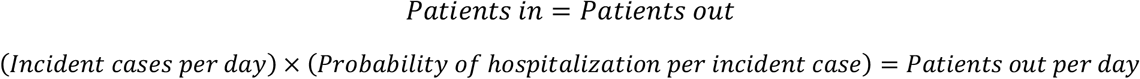

Knowing the rate of patients leaving the hospital (when at capacity with COVID-19 patients), and the expected probability of hospitalization per case, we can solve for incident cases per day:

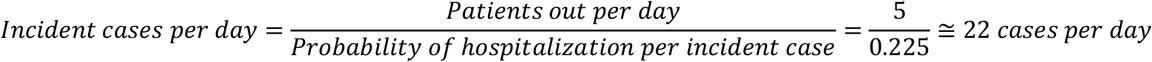

Therefore, a healthcare system can sustain a maximum of approximately 22 new cases per day with the hospital at full-capacity for COVID-19 patients. This would result in 22 patients diagnosed with COVID-19 per day, of which 5 will be admitted to hospital, and 5 COVID-19 patients leaving the hospital either through discharge or death.

### Extension to other resources

In this example we have demonstrated the application using acute care hospitalizations. This methodology can be applied to any resource of interest. In our online tool we apply it to acute care beds, critical care beds, and mechanical ventilators. For each resource one must know the average length-of-use per admitted COVID-19 patient (for the cases). It is important to use the ***average*** duration of use and not the median. It is also important to separate lengths of use as mutually exclusive and non-mutually exclusive when appropriate. For example, a patient is admitted to hospital, spends the first 5 days in acute care, 6 days in critical care (no intubation), followed by another 6 days in critical care intubated, another 2 days in critical care (no intubation), and 5 more days in acute care. In this example, the length-of-stay in acute care is 10 days, the length-of-stay in critical care is 14 days, and length of mechanical ventilation is 6 days. *In our tool it is assumed that a ventilated patient is de facto in critical care*. Therefore the number of ventilators should not exceed the number of critical care beds.

## References

1. Zhou F, Yu T, Du R, et al. Clinical course and risk factors for mortality of adult inpatients with COVID-19 in Wuhan, China: a retrospective cohort study. Lancet. 2020.

2. World Health Organization. Coronavirus disease 2019 (COVID-19) Situation Report – 61. https://www.who.int/docs/default-source/coronaviruse/situation-reports/20200321-sitrep-61-covid-19.pdf?sfvrsn=6aa18912_2. Published March 21, 2020. Accessed March 22, 2020.

3. Grasselli G, Pesenti A, Cecconi M. Critical Care Utilization for the COVID-19 Outbreak in Lombardy, Italy: Early Experience and Forecast During an Emergency Response. JAMA. 2020.

4. Dong J, Perry O. Queueing Models for Patient-Flow Dynamics in Inpatient Wards. Operations Research. 2020;68(1):250–275.

5. Wang D, Hu B, Hu C, et al. Clinical Characteristics of 138 Hospitalized Patients With 2019 Novel Coronavirus-Infected Pneumonia in Wuhan, China. JAMA. 2020.

6. CDC COVID-19 Response Team. Severe Outcomes Among Patients with Coronavirus Disease 2019 (COVID-19) — United States, February 12–March 16, 2020. MMWR Morb Mortal Wkly Rep. 2020.

